# Is it possible to influence Pusher Syndrome with Tilted Reality? A series of four cases

**DOI:** 10.64898/2025.12.27.25340867

**Authors:** Sofia Müller-Wöhrstein, Hans-Otto Karnath

## Abstract

Pusher syndrome is a disorder of postural control, characterized by an altered perception of upright body orientation. While visual verticality perception remains intact, patients incorrectly perceive their tilted body posture as upright, resulting in active resistance to posture correction. This perceptual mismatch offers a potential target for therapeutic interventions. We evaluated a newly developed *Tilted Reality Device* (TRD) designed to recalibrate body verticality perception by subtly tilting the patient’s real-time visual environment towards the ipsilesional side in a series of four patients with pusher syndrome. We implemented a three-phase experimental design: pre-manipulation (normal view), manipulation (TRD tilted 20° ipsilesionally), and post-manipulation (normal view). Passive body tilts were performed while we recorded body orientation. Fifteen healthy older adults served as controls. The behavioral changes were assessed via tilt angle analysis and were statistically compared using Crawford-Garthwaite Bayesian methods. Two of the four patients showed reduced resistance to ipsilesional body tilts during the TRD manipulation. Two patients also demonstrated an aftereffect. Our findings only partly confirmed our expectations; possible limitations are discussed. Further research is needed to evaluate our TRD in its potential treatment effects in pusher syndrome and better understand the mechanisms involved in the recalibration of upright body orientation perception in patients with pusher syndrome.

## Introduction

Pusher syndrome is a disorder of postural control (Davies, 1985; Karnath et al., 2000b) that occurs in approximately 12.5% of hemiparetic stroke patients (Abe et al., 2012; Dai et al., 2022; Pedersen et al., 1996). It is characterized clinically by three criteria: i) patients have a spontaneous body posture that is inclined towards the contralesional side of the body; ii) they use their unaffected limbs (arms and/or legs) to push themselves further toward the contralesional paretic side of the body, so that they eventually fall to that side; and iii) they resist external posture correction by abducting and extending their arms and/or legs, thus blocking passive movement (Davies, 1985; Karnath, 2007; Karnath et al., 2001). These three criteria are also part of the diagnostically used Scale for Contraversive Pushing (SCP; Karnath et al., 2000b). The cause of pusher syndrome is usually a hemorrhagic stroke, slightly more common in right-hemispheric than left-hemispheric strokes (Abe et al., 2012; Karnath et al., 2000a). The most relevant lesion site is thought to be the posterolateral thalamus (Karnath et al., 2000a; Karnath et al., 2005), while cases of pusher syndrome following cortical strokes have also been described (Johannsen, Brötz, et al., 2006; van der Waal et al., 2022). A recent study has revealed a white matter disconnection between the thalamus and the cortex (Rosenzopf et al., 2023), which might explain the previous findings of both thalamic and cortical lesions involved in pusher syndrome.

Pusher syndrome is caused by a faulty perception of onés own body position in space (Karnath et al., 2000b). Patients in the acute phase of stroke perceive themselves as being upright (that is in line with the earth’s vertical axis), although they are actually tilted by an average of ∼18° toward their ipsilesional side (Bergmann et al., 2016; Karnath et al., 2000b). Contrary to this false perception of vertical body orientation, the correct alignment of visually presented vertical structures is not impaired in pusher patients (Baier et al., 2012; Johannsen, Fruhmann Berger, et al., 2006; Karnath et al., 2000b). This leads to a mismatch in the perception of verticality, whereby the perception of visual input is intact, but the perception of body orientation is disturbed. This mismatch is assumed to be the underlying pathological mechanism of pusher syndrome (Karnath et al., 2000b). At the same time, this mismatch can be used as a starting point for therapeutic approaches. Since visual verticality perception is intact, it can be used to train and improve the faulty perception of body verticality in pusher syndrome patients. This therapeutic principle is used by the Visual Feedback Training (VFT; Brötz et al., 2004; Karnath & Brötz, 2003), in which visual aids – such as the therapist’s vertically aligned arm – are used to actively help the patient consciously achieve the correct upright body posture. Another approach is to use the undisturbed visual sense to convey false information about the visual environment in relation to the patient’s own body orientation to the brain, so that this could lead to an “automatic” correction of the patient’s posture. This functional principle has already led to an improvement in symptoms of pusher syndrome in a single-case study by Nestmann et al. (2022). The authors used an artificial virtual reality (VR) environment and manipulated the visual scene by tilting the horizon. Since using the real visual environment is closer to everyday life, more cost-effective, and can also be integrated into physiotherapy sessions, we have developed a *Tilted Reality Device* (Wöhrstein et al., 2025). Patients wear a head-mounted display with cameras that record their actual surroundings. The TRD thus shows patients real-time visual footage of their actual environment. What makes the TRD special is that the mounted cameras can be rotated so that the images of the visual environment can also be presented to the patient tilted on roll plane. We have found no contraindications for the use of the TRD in older individuals (Wöhrstein et al., 2025). Therefore, the aim of this study is to evaluate the therapeutic use of the TRD in a first case-series of patients with pusher syndrome.

## Methods and Materials

### Pusher Patients

Patients with pusher syndrome following unilateral brain damage, consecutively admitted to the Center of Neurology at Tübingen University Hospital or to the Clinic for Neurology and Early Rehabilitation at Reutlingen District Hospital, were included in the study. Structural imaging was acquired by computed tomography (CT) and/or magnetic resonance (MR) imaging as part of the clinical routine procedure carried out in the acute phase at stroke-onset. Patients in whom imaging did not reveal any clear lesions were not included. Inclusion was based on meeting the following behavioral criteria: Each of the three items of the Scale for Contraversive Pushing (Baccini et al., 2008; Karnath et al., 2007; Karnath et al., 2000b) had to be scored as a total of 1 by a trained physiotherapist or occupational therapist. If the patient was unable to stand, the seated measurement necessarily had to be scored as 1. Our case series included four consecutively admitted patients who met these inclusion criteria for pusher syndrome. Demographic and clinical details are given in Table 1; the brain imaging data in Fig 1.

**Table 1.** Demographic and clinical characteristics for the pusher syndrome patients.

|  | **P1** | **P2** | **P3** | **P4** |
| --- | --- | --- | --- | --- |
| Age (years) | in his 70's | in her 80's | in his 70's | in her 70's |
| Sex | male | female | male | female |
| Lesion side | LH | RH | RH | RH |
| Hemiparesis | Right arm 2/5 | Left arm 0/5 | Left arm 2/5 | Left arm 4/5 |
|  | Right leg 2/5 | Left leg 2/5 | Left leg 2/5 | Left leg 3/5 |
| SCP_A sitting | 1 | 1 | 1 | 1 |
| SCP_A standing | n.p. | 1 | n.p. | n.p. |
| SCP_B sitting | 1 | 1 | 1 | 1 |
| SCP_B standing | n.p. | 1 | n.p. | n.p. |
| SCP_C sitting | 1 | 1 | 1 | 1 |
| SCP_C standing | n.p. | 1 | n.p. | n.p. |
| Time between symptom onset and experimental testing (days) | 21 | 19 | 45 | 41 |

*Note.* The degree of hemiparesis of the upper and lower limbs was scored with the usual clinical ordinal scale, where 0 is no trace of movement and 5 is normal movement. The values of the Scale for Contraversive Pushing (SCP; Karnath et al., 2007; Karnath et al., 2000b) correspond to the following items: SCP\_A = spontaneous body posture with possible values 0, 0.25, 0.75, 1; SCP\_B = use of non-paretic extremities (abduction and extension) with possible values 0, 0.5, 1; SCP\_C = behavior during passive correction of body posture with possible values 0, 1. Each item is tested during sitting as well as during standing if possible. Exact age is not specified here due to medRxiv guidelines. n.p. = testing not possible; LH = left hemisphere; RH = right hemisphere.

**Fig 1.**
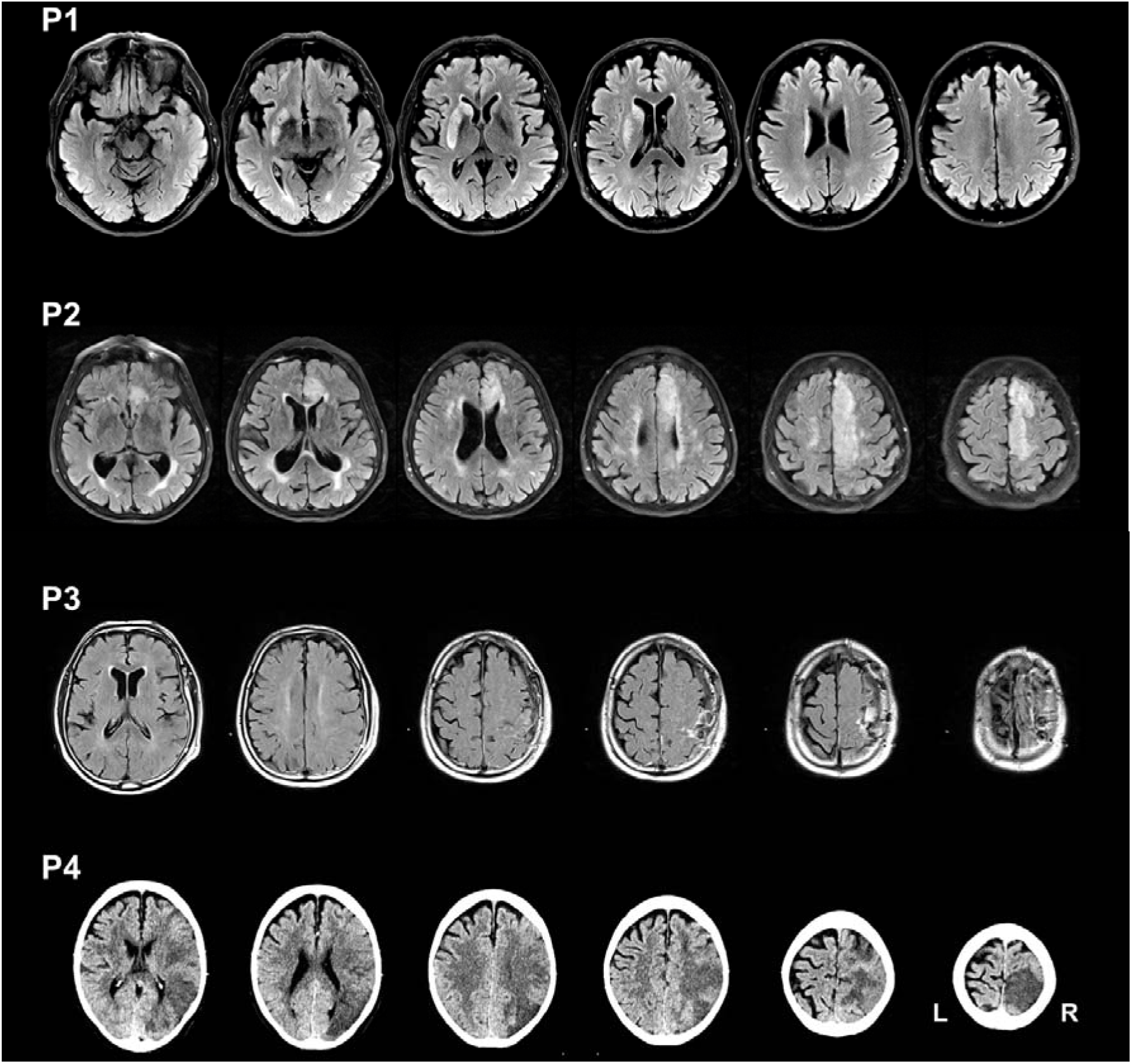
Acute imaging scans of all four patients, taken within the first 72 hours after symptom onset. The slices shown vary from patient to patient in order to best visualize the lesion. For patient **P1**, **P2**, and **P3** FLAIR MR scans were taken; for patient **P4** a CT scan was available.

Patient P1 was a man in his 70’s who initially suffered a left-hemispheric ischemic stroke followed by a hemorrhagic stroke in the left basal ganglia. At the time of testing, he presented with right-sided hemiparesis and facial nerve palsy as well as mild aphasia. Patient P2 was a woman in her 80’s who suffered a right-hemispheric ischemic stroke in the anterior cerebral artery territory. She presented with left-sided hemiparesis, psychomotor deceleration and language impoverishment. Patient P3 was a man in his 70’s who suffered from a right-hemispheric fronto-parietal hemorrhaged meningothelial meningioma. Six weeks after his osteoplastic trepanation with removal of the meningioma, he presented with left-sided hemiparesis and coordination disorders. Finally, patient P4 was a woman in her 70’s who suffered a right-hemispheric ischemic stroke in the middle cerebral artery territory. She presented with left-sided brachiofacial hemiparesis and dysarthria. In all four patients, formal neuropsychological assessment could not be performed at the time of experimental testing because of their severe health condition.

All participants, both patients and healthy controls (see next section), provided their written informed consent to participate in the study prior to its commencement. The study was conducted in accordance with the Code of Ethics of the World Medical Association (revised version of the 1964 Declaration of Helsinki). In addition, patients gave their written consent to publish medical details as well as imaging data for scientific purposes. The study was approved by the Ethics Committee of the Faculty of Medicine of the University of Tübingen, Germany (814/2021BO2).

### Healthy Controls

In order to obtain a baseline of healthy individuals’ behavior for the experimental manipulations, we recruited 15 older adults (7 females) over the age of 60 as healthy control participants. The average age of the control group was 68.5 years (62 to 85 years, SD = 5.8). None of the healthy controls had a history of vestibular or oculomotor abnormalities and all were free from any neurological or psychiatric disorders. Healthy controls were financially compensated for their participation.

### Experimental Procedure

The study was pre-registered (aspredicted.org #91254). The procedure started with an assessment of the SCP immediately prior to the start of the actual experiment. During the experiment, patients sat on an examination couch with their feet hanging freely and were supported by a trained physiotherapist or occupational therapist who knelt behind the patient on the couch. The physiotherapist or occupational therapist also assisted throughout the experiment, ensuring that the patient was safe and not at risk of falling. Opposite the patient’s sitting position was either a window with many visible window frames or, if the patient was facing a wall, the wall was covered with vertical stripes for the patient to look at. This ensured that the patient could always see vertical structures in his or her visual environment. The room was always brightly illuminated.

At the beginning of the experiment, the experimenter put the TRD on the patient and made sure he or she was comfortable with it (for technical details see Wöhrstein et al., 2025). The patient was instructed to keep his or her head upright, eyes open, and to look straight ahead during the experiment. Upright head position was visually controlled by the experimenter. The patient’s hands were placed loosely in his or her lap (without instructions to keep the hands on the lap during the experiment). The study comprised three consecutive experimental conditions: (i) measurement without tilt (0°) of the TRD (pre-manipulation condition *A*), (ii) measurement with the TRD tilted 20° to the ipsilesional side (manipulation condition *B*), and (iii) measurement, again without tilt (0°) of the TRD (post-manipulation condition *A’*). The center of rotation was aligned with the patient’s eye level. Under each of the three conditions, five cycles of passive body movements were performed in roll plane: the physiotherapist or occupational therapist slowly tilted the patient to one side and then to the other, always starting with the patient’s contralesional, paretic side. Each patient was verbally instructed to “allow the movement”, in line with the SCP instructions (Karnath et al., 2007). Towards the contralesional side, the passive tilt movement was stopped by the therapist at a tilt angle of ∼35-40°. Towards the ipsilesional side, the tilt movement was stopped as soon as the patient used his or her unparetic, ipsilesional arm to resist and block the experimenter’s movement. Such active resistance to passive movements is a very typical behavior of patients with pusher syndrome (Davies, 1985; Karnath et al., 2001) and is also part of the SCP criteria (Baccini et al., 2008; Karnath et al., 2007; Karnath et al., 2000b). Therefore, it was chosen as stop criterion for the experimental manipulation. Between each tilt to one side, the experimenter ensured that both hands were resting loosely on the patient’s lap again and that the TRD remained comfortable for the patient.

For healthy control participants, the procedure was similar to that for patients, but without SCP assessment. The passive tilt movements were stopped by the experimenter on both sides at a tilt angle of ∼35-40°. The direction of the first body tilt as well as the tilt direction of the TRD were randomized for all participants to avoid possible biases. Seven participants began with a body tilt to the left and eight with a body tilt to the right. The tilt direction of the TRD was matched to the direction of the first body tilt: if the first body tilt was to the left, the TRD was tilted to the right in condition *B*; if the first body tilt was to the right, the TRD was tilted to the left in condition *B*. This was based on the logic of patient measurements, in which the tilt direction of the TRD was always set to the opposite side of the initial body tilt.

The longitudinal body axis of all patients and healthy controls in the three experimental conditions was recorded with markerless motion capture, using two video cameras (Sony HDR-CX240 HD Flash Memory Camcorder) with a resolution of 1920 x 1080p and a frame rate of 50 frames per second. The two cameras were positioned 20° to the front left and front right of the participant.

### Data Analysis

Video data were anonymized by wearing the TRD, which covered the eyes and nose of the participants and by removing the audio from the video. Initial extraction of motion tracking keypoints from the video recordings was performed by an external company (Subsequent GmbH, Konstanz, Germany), using a deep learning algorithm based on a multi-stage convolutional neural network. Individual 3D joint coordinates for each joint point in each video frame were computed and extracted from the two video recordings per patient or healthy control (for technical details see Barzyk et al., 2024). These joint coordinates were used to perform the following analysis.

We manually selected the corresponding video frames for each of the three experimental conditions (conditions *A*, *B*, and *A’*) and for the upright starting position. From the coordinates of the hip center and the neck, we formed a vector that indicates the respective body position of the patient (cf. Fig. 2). Since we had multiple measurements for each of the three experimental conditions, the individual vectors were averaged to form one mean vector for each condition and each tilt side. Using these mean vectors, we then calculated the angles between the baseline vector (that is the upright starting position) and the vectors of each condition on each side of the body using the arccosine function. This resulted in six tilt angles for each participant, representing the change in (upright) body orientation: *condition A_left*, *condition A_right*, *condition B_left*, *condition B_right*, *condition A’_left*, and *condition A’_right*. For the healthy controls, in addition, the angles of all control participants were averaged per condition to obtain a mean control angle for each of the three experimental conditions for both tilt sides.

**Fig 2.**
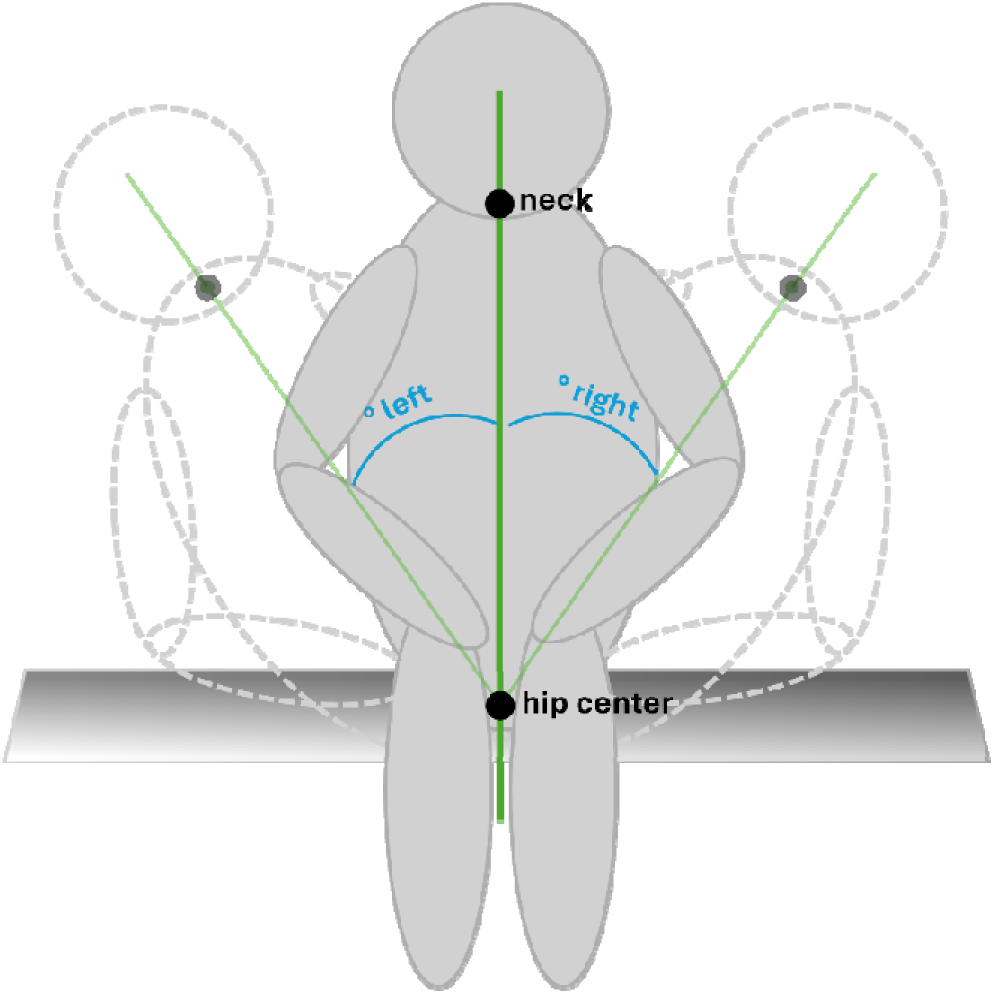
Schematic representation of the calculation of vectors and angles of body position for one trial. The bold green line represents the baseline longitudinal body axis, including the joint points at hip center and neck. The transparent green lines represent body tilts to the left and right, respectively. The respective angles of body tilt are shown in blue.

### Statistical Analysis

We used the Crawford-Garthwaite Bayesian test for single-case analysis (Crawford & Garthwaite, 2007) to determine the individual patients’ behavior in comparison to the healthy control group. We used the overall mean tilt angle of all healthy controls, regardless of tilt side or experimental condition, as baseline to which the individual patients’ tilt angles per condition were compared to. This was done using the *crawford.test()* function from the *psycho* package in *R* (Makowski, 2018). Additionally, we used the revised standardized difference test (Crawford & Garthwaite, 2005) to detect possible intra-individual improvements in a single patient between ipsilesional tilts in conditions *A* and *B* as well as conditions *A* and *A’*. The mean differences between the corresponding conditions, regardless of tilt side, in healthy controls were used as reference values. For implementation of this test, we used the *RSDT()* function from the *singcar* package in *R* (Rittmo & McIntosh, 2021). All data processing and analyses mentioned were performed using *R Studio* (R Version 4.4.0; Posit Software, 2024).

## Results

### Healthy Controls

None of the healthy control participants resisted the passive body tilts in any of the three conditions. The average tilt angle across all healthy controls, conditions, and tilt directions was 37.61° (*SD* = 11.27°). The mean angles for all three experimental conditions and tilt sides from the group of healthy controls are shown in Table 2.

**Table 2.** Mean angles including standard deviations for the passive body tilts for all experimental conditions and tilt sides in healthy controls (N = 15).

| Condition | Mean<br>tilt angle | Standard Deviation<br>tilt angle |
| --- | --- | --- |
| *A_left* | $37.54^{\circ}$ | $10.25^{\circ}$ |
| *A_right* | $34.44^{\circ}$ | $10.12^{\circ}$ |
| *A_mean* | $35.99^{\circ}$ | $10.13^{\circ}$ |
| *B_left* | $39.45^{\circ}$ | $12.67^{\circ}$ |
| *B_right* | $34.81^{\circ}$ | $11.30^{\circ}$ |
| *B_mean* | $37.13^{\circ}$ | $12.03^{\circ}$ |
| *A'_left* | $41.50^{\circ}$ | $10.88^{\circ}$ |
| *A'_right* | $37.90^{\circ}$ | $12.39^{\circ}$ |
| *A'_mean* | $39.70^{\circ}$ | $11.60^{\circ}$ |

*Note.* *A* stands for the pre-manipulation condition *A* with the Tilted Reality Device (TRD) without tilt ( $0^{\circ}$ ), *B* for the manipulation condition *B* with the TRD tilted $20^{\circ}$ toward the ipsilesional side of the patient, and *A'* for the post-manipulation condition *A'* without tilt ( $0^{\circ}$ ) of the TRD. For each of the three conditions, there were body tilts to the left and right.

### Pusher Patients

Results of the Crawford-Garthwaite Bayesian test for single-case analysis of all patients under all experimental conditions are listed in Table 3 and graphically represented in Fig 3.

**Table 3.**
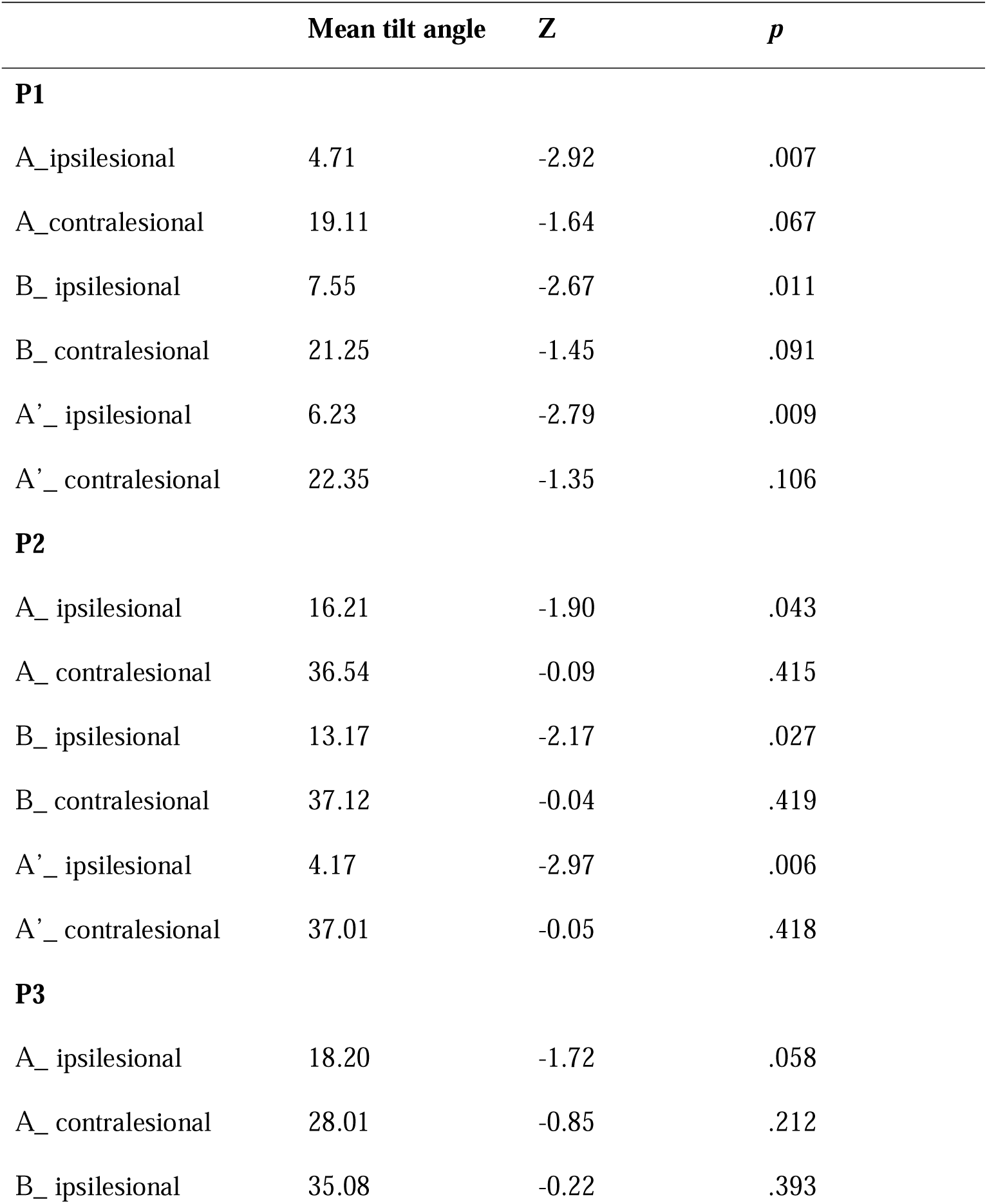
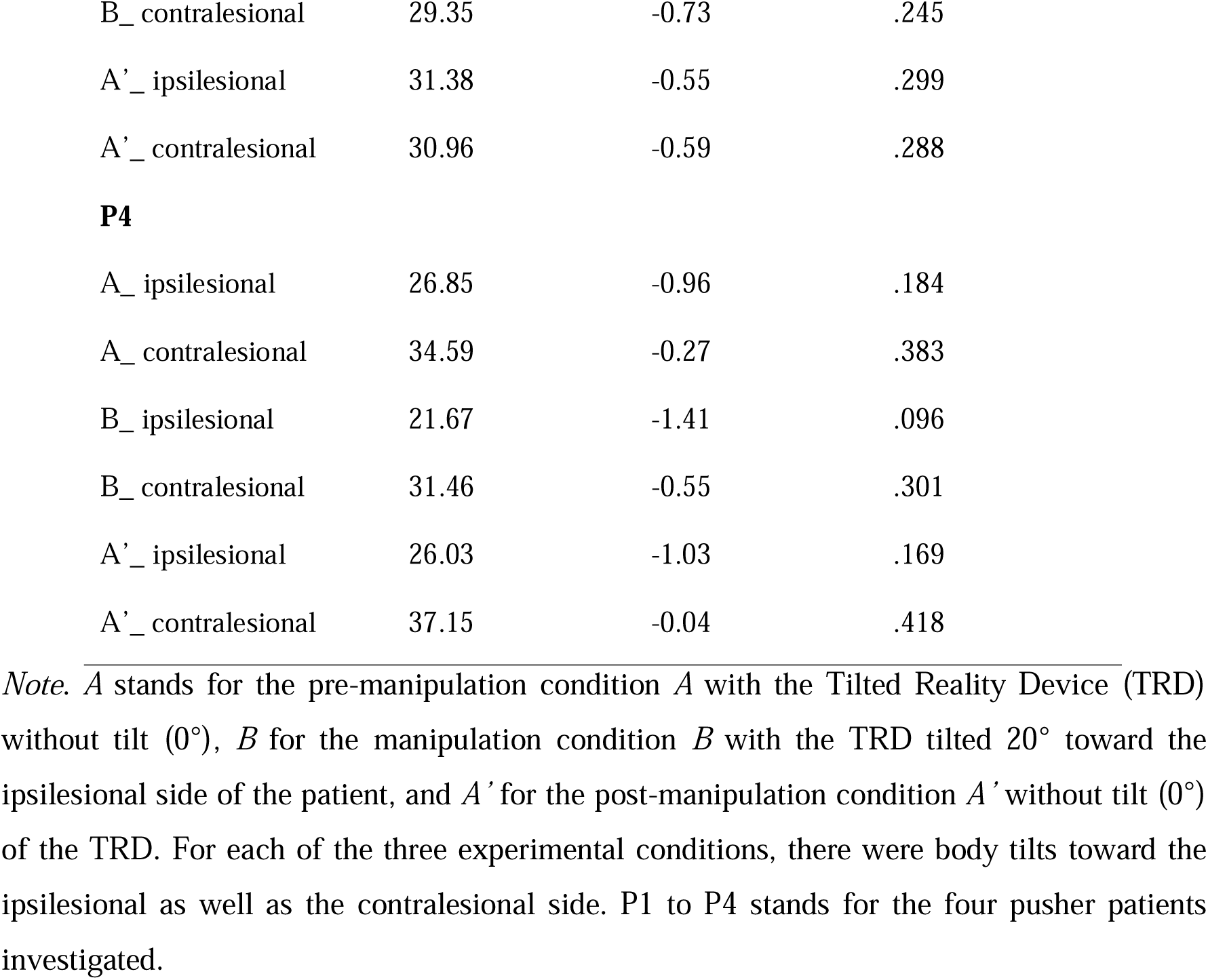
Results from the Crawford-Garthwaite Bayesian test for single-case analysis for all four pusher patients (P1 to P4) under all experimental conditions.

|  | Mean tilt angle | Z | *p* |
| --- | --- | --- | --- |
| **P1** |  |  |  |
| A_ipsilesional | 4.71 | -2.92 | .007 |
| A_contralesional | 19.11 | -1.64 | .067 |
| B_ipsilesional | 7.55 | -2.67 | .011 |
| B_contralesional | 21.25 | -1.45 | .091 |
| A'_ ipsilesional | 6.23 | -2.79 | .009 |
| A'_ contralesional | 22.35 | -1.35 | .106 |
| **P2** |  |  |  |
| A_ipsilesional | 16.21 | -1.90 | .043 |
| A_contralesional | 36.54 | -0.09 | .415 |
| B_ipsilesional | 13.17 | -2.17 | .027 |
| B_contralesional | 37.12 | -0.04 | .419 |
| A'_ ipsilesional | 4.17 | -2.97 | .006 |
| A'_ contralesional | 37.01 | -0.05 | .418 |
| **P3** |  |  |  |
| A_ipsilesional | 18.20 | -1.72 | .058 |
| A_contralesional | 28.01 | -0.85 | .212 |
| B_ipsilesional | 35.08 | -0.22 | .393 |
| B_ contralesional | 29.35 | -0.73 | .245 |
| A'_ ipsilesional | 31.38 | -0.55 | .299 |
| A'_ contralesional | 30.96 | -0.59 | .288 |
| **P4** |  |  |  |
| A_ ipsilesional | 26.85 | -0.96 | .184 |
| A_ contralesional | 34.59 | -0.27 | .383 |
| B_ ipsilesional | 21.67 | -1.41 | .096 |
| B_ contralesional | 31.46 | -0.55 | .301 |
| A'_ ipsilesional | 26.03 | -1.03 | .169 |
| A'_ contralesional | 37.15 | -0.04 | .418 |

*Note.* *A* stands for the pre-manipulation condition *A* with the Tilted Reality Device (TRD) without tilt ( $0^\circ$ ), *B* for the manipulation condition *B* with the TRD tilted $20^\circ$ toward the ipsilesional side of the patient, and *A'* for the post-manipulation condition *A'* without tilt ( $0^\circ$ ) of the TRD. For each of the three experimental conditions, there were body tilts toward the ipsilesional as well as the contralesional side. P1 to P4 stands for the four pusher patients investigated.

**Fig 3.**
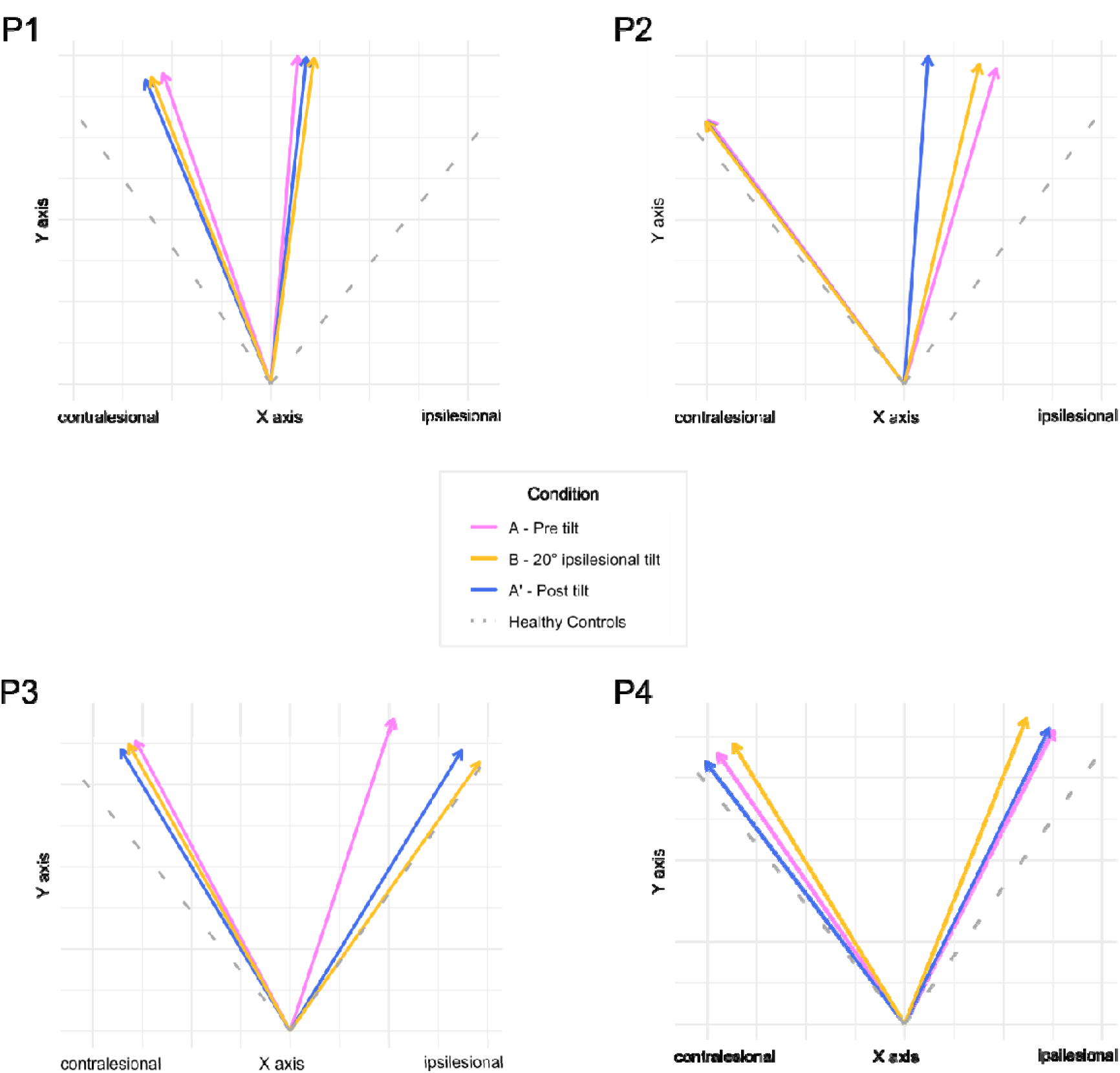
Extent of tilt to the ipsilesional and contralesional side of the body of the four pusher patients. Arrows represent the individual average tilt for each color-coded condition, the dotted lines represent the mean tilt for healthy control subjects.

#### Pre-manipulation condition *A*

As expected, none of the pusher patients showed active resistance to passive body tilts toward their contralesional side in the pre-manipulation condition *A* (Fig. 3). In all four pusher patients, we found no statistically significant differences compared to healthy controls in the tilt angles toward the contralesional side (cf. Table 3). However, they all resisted passive body tilts toward their ipsilesional side by abducting and extending their non-paretic arm and hand from their lap to the hospital couch, thereby stopping the passive movement. Accordingly, we found smaller body tilts in all four patients with pusher syndrome when it came to body tilts towards their ipsilesional side (Fig. 3). In pusher patients P1 and P2 these tilts were statistically significantly smaller to healthy controls, and numerically, but not statistically significantly, smaller in patients P3 and P4 (cf. Table 3).

#### Manipulation condition *B*

We first investigated whether or not wearing the TRD changes pusher patients’ active resistance to passive movements toward the ipsilesional side to a “normal” level, that is a level that is also observed in healthy controls. In patients P1 and P2, we still found a statistically significant difference in tilt angles compared to healthy controls, while this was not the case for patients P3 and P4 (cf. Table 3). For patients P3 and P4, we still found no significant difference in their ipsilesional body tilts compared to healthy controls (cf. Table 3). However, in patient P3, the ipsilesional tilt angle was numerically comparable to that of healthy controls, while, in patient P4, the ipsilesional tilt angle was still numerically smaller than in healthy controls but did not reach statistical significance (cf. Table 3).

Next, we investigated whether or not wearing the TRD reduces pusher patients’ active resistance to passive movements toward the ipsilesional side. Therefore, we compared the difference between ipsilesional tilt angles under conditions *A* and *B* for each individual pusher patient with the difference observed in healthy controls, using the revised standardized difference test. In patient P1, we found that only 3.1% of healthy controls showed a larger difference in their tilt angles between conditions *A* and *B*. Nevertheless, the difference between conditions *A* and *B* failed − even if only narrowly − statistical significance when compared to the differences observed in healthy controls (*t*(14) = 2.03, *p* = .062). In patient P2, 45.0% of healthy controls showed a difference as large or larger in their tilt angles between conditions *A* and *B.* The difference between conditions *A* and *B* was not statistically significantly larger than the differences observed in healthy controls (*t*(14) = 0.13, *p* = .900). In patient P3, only 0.01% of healthy controls showed a larger difference in their tilt angles between conditions *A* and *B*; the difference between ipsilesional tilt angles in conditions *A* and *B* was statistically significantly larger than the differences observed in healthy controls (*t*(14) = 4.92, *p* = < .001), indicating a clear positive effect of the TRD. Lastly, in patient P4, 11.7% of healthy controls showed a difference as large or larger in their tilt angles between conditions *A* and *B*. The difference between conditions *A* and *B* did not reach statistical significance when compared to the differences observed in healthy controls (*t*(14) = 1.25, *p* = .233).

#### Post-manipulation condition *A’*

As a final step, we investigated whether or not wearing the TRD tilted toward the ipsilesional side had an aftereffect in patients with pusher syndrome, that is, whether it changed their pathological behavior to a “normal” level once the patients regained a vertically aligned view. Here, too, we compared the data from the four individual pusher patients with that from healthy controls. In patients P1 and P2, we did not find any reduction in their pathological behavior compared to healthy controls; the difference was still statistically significant in tilt angles (cf. Table 3). In patients P3 and P4, the ipsilesional tilt angles did not reach statistical significance (cf. Table 3), even though they were still numerically smaller compared to healthy controls.

Lastly, we also investigated whether or not wearing the TRD tilted toward the ipsilesional side had a positive aftereffect in individual patients with pusher syndrome, that is, whether it reduced their active resistance to passive movements toward the ipsilesional side once the patients regained a vertically aligned view. For this purpose, we compared the difference between ipsilesional tilt angles in conditions *A* and *A’* for each individual pusher patients, using the revised standardized difference test. In patient P1, 34.1% of healthy controls showed a larger difference in their tilt angles between conditions *A* and *A’*. The difference between conditions *A* and *A’* was not statistically significantly larger than the differences observed in healthy controls (*t*(14) = 0.42, *p* = .682). In patient P2, only 2.0% of healthy controls showed a difference as large or larger in their tilt angles between conditions *A* and *A’*. The difference between conditions *A* and *A’* was statistically significantly larger than the differences observed in healthy controls (*t*(14) = 2.26, *p* = .040). The ipsilesional tilt angle in the post-manipulation condition *A’* was significantly smaller than in the pre-manipulation condition *A*, suggesting that the TRD had an effect, but not in the expected direction. In patient P3, only 2.6% of healthy controls showed a larger difference in their tilt angles between conditions *A* and *B*. The difference between ipsilesional tilt angles in conditions *A* and *A’* only narrowly failed statistical significance when compared to the differences observed in healthy controls (*t*(14) = 2.12, *p* = .052); numerically the tilt angle was larger in the post-manipulation condition *A’* than in the pre-manipulation condition *A* and thus indicated a trend towards a positive aftereffect of the TRD. Lastly, in patient P4, 28.9% of healthy controls showed a difference as large or larger in their tilt angles between conditions *A* and *A’*. The difference between conditions *A* and *A’* was not statistically significantly larger than the differences observed in healthy controls (*t*(14) = 0.57, *p* = .589).

## Discussion

We evaluated the therapeutic use of a new TRD in a case-series of four patients with pusher syndrome. Our results partly confirmed our expectations. Two of our patients (P1 and P3) showed a positive effect in that their active resistance to passive ipsilesional body tilts was reduced during the manipulation with the TRD if the visual surroundings were tilted to the ipsilesional side. This effect was statistically significant in P3 and numerically present in the same direction in P1. In contrast, we found no such effect in pusher patients P2 and P4.

In detail, patient P1 differed from our group of healthy control subjects in terms of the extent of his tilt angles in all three experimental conditions, suggesting that his pathological behavior was not reduced to a “normal” level of healthy controls by the use of the TRD. He showed a small numerical improvement between the baseline measurement and the tilt manipulation condition, suggesting that the TRD had a small effect during the tilted view. Patient P2 showed the same pattern as patient P1 in terms of group comparison with regard to the lack of normalization of her pathological behavior. At the individual level, no effect of the TRD was observed for P2 between the baseline measurement and the tilt manipulation condition. In contrast to these two patients, patients P3 and P4 did not differ from our group of healthy controls in terms of the extent of their tilt angles in all three experimental conditions. At the individual level, patient P3 showed an improvement between the baseline measurement and the tilted manipulation condition in terms of less resistance to passive body movements. Patient P4 showed no improvement in terms of the extent of her tilt angles on an individual level between the baseline measurement and the tilt manipulation condition.

With respect to aftereffects between the baseline measurement and the post-manipulation condition, we observed that the TRD had an effect in two of our patients (P2 and P3). Patient P3 allowed numerically larger ipsilesional body tilts, while patient P2 surprisingly allowed only less movement, which was in the opposite direction to what was expected. We did not observe any such aftereffects in pusher patients P1 and P4 who did not show a numerical or statistical improvement between the baseline measurement and the post-manipulation condition.

One possible explanation for the heterogenous results could be the variability between the pusher patients included. Although we aimed to include patients with similar behavioral profiles, our findings revealed that two of the four pusher patients (patients P3 and P4) were statistically closer to normal, non-pathological behavior in terms of the extent of their body tilt angles at the start of the experiment. This suggests that their degree of pathological behavior was presumably less pronounced from the onset, potentially skewing our outcomes. Furthermore, the four patients varied in terms of lesion location and size, etiology, and stage of recovery. Recovery stage is particularly crucial, as it can significantly influence pusher patients’ pathological behavior. The two pusher patients who did not demonstrate significant deviations from the control group at the start of the experiment (patients P3 and P4) were already in the early rehabilitation phase, while the other two patients were still in the acute/subacute phase of the stroke. This difference in the stage of recovery might indicate that they had already benefited from physiotherapy interventions, including Visual Feedback Training (VFT), which has been shown to help pusher patients in counteracting their pathological posture (Brötz et al., 2004; Yang et al., 2015). It might be plausible that such therapies have already taught these patients effective compensation strategies to combat their pathological behavior, which they might have also used during our experiment, leading to the observed body tilt angles that were closer to normal values. Beyond, with regard to lesion characteristics, our case-series of four patients with pusher syndrome did not include any patients with thalamic hemorrhagic lesions, particularly in the ventral posterior and lateral posterior nuclei of the posterolateral thalamus, which are typically associated with pusher syndrome (Karnath et al., 2000a; Rosenzopf et al., 2023). These lesions are assumed to affect the neural pathways crucial for processing trunk position and upright body posture (Karnath et al., 2000a; Mittelstaedt, 1992; Mittelstaedt, 1998), and their absence in our case-series may have contributed to the heterogeneity in responses to the TRD.

A further source that might have contributed to our mixed results is the technique used to manipulate the behavior of the patients. Compared to the study by Nestmann et al. (2022), in which a single patient with pusher syndrome achieved significant improvement through exposure to a tilted VR scene, the present study used a new Tilted Reality Device (TRD) instead of VR. It is possible that the immersive and multisensory nature of VR provides a more compelling stimulus for behavioral changes compared to the TRD. However, it is likewise possible that pusher patients might require a longer exposure to the tilted environment provided by the TRD for its effects to be reflected in our outcome measures. In our present experiment, we started the cycles of passive body movements immediately after putting on the TRD in the non-tilted position on the patient. This means that the patients’ exposure to the tilted environment lasted only as long as the tilt cycles in experimental condition *B*, which was only a few minutes. The patients therefore did not have much time to adjust to the new visual environment. It is possible that their exposure to the tilted environment was too short as a result. The learning process associated with adapting to new sensory stimuli, such as visual stimuli, particularly in individuals with altered perception such as those with pusher syndrome, may not result in immediate or observable changes in behavior. Prolonged exposure to the tilted environment could offer patients more opportunities to develop and consolidate functional compensation strategies that are essential for restoring their postural control. This could be crucial for reducing the mismatch between visual and vestibular input, which acts as the pathological mechanism in pusher syndrome (Karnath et al., 2000b).

To summarize, at this point, further research is needed to evaluate our TRD in its potential treatment effects in pusher syndrome and better understand the mechanisms involved in the recalibration of upright body orientation perception in patients with pusher syndrome. Larger studies are needed to prove or disprove our hypothesis and potentially identify patient subgroups who are most likely to benefit from treatment with the TRD. Understanding how variations in exposure duration along with other factors, such as individual cognitive and physical abilities, affect the rehabilitation and intervention outcomes is essential for developing optimized treatment protocols.

## Acknowledgements

We would like to thank all patients for their participation in our study. Additionally, we also like to thank all physiotherapists and occupational therapists who assisted us during recruiting and testing of the patients. Finally, we would like to thank Tim Dreßler for his help with data acquisition as well as Dr. Peter Müller-Wöhrstein for his support with data analysis.

## Author Contributions

S.M-W. collected the data, performed all analyses, and wrote the first version of the manuscript. H-O.K. provided resources, planned and supervised the study and commented on previous versions of the manuscript. All authors read and approved the final version of the manuscript.

## Data Availability

The data that support the findings of this study are available from the corresponding author upon reasonable request.

## Abbreviations

*SCP*: Scale for Contraversive Pushing
*TRD*: Tilted Reality Device
*VR*: virtual reality

## References

Abe, H., Kondo, T., Oouchida, Y., Suzukamo, Y., Fujiwara, S., & Izumi, S. I. (2012). Prevalence and length of recovery of pusher syndrome based on cerebral hemispheric lesion side in patients with acute stroke. Stroke, 43(6), 1654–1656. 10.1161/STROKEAHA.111.638379

Baccini, M., Paci, M., Nannetti, L., Biricolti, C., & Rinaldi, L. A. (2008). Scale for contraversive pushing: cutoff scores for diagnosing “pusher behavior” and construct validity. Physical Therapy, 88(8), 947–955. 10.2522/ptj.20070179

Baier, B., Janzen, J., Müller-Forell, W., Fechir, M., Müller, N., & Dieterich, M. (2012). Pusher syndrome: its cortical correlate. Journal of Neurology, 259(2), 277–283. 10.1007/s00415-011-6173-z

Barzyk, P., Boden, A.-S., Howaldt, J., Stürner, J., Zimmermann, P., Seebacher, D., Liepert, J., Stein, M., Gruber, M., & Schwenk, M. (2024). Steps to Facilitate the Use of Clinical Gait Analysis in Stroke Patients: The Validation of a Single 2D RGB Smartphone Video-Based System for Gait Analysis. Sensors, 24(23), 7819. 10.3390/s24237819

Bergmann, J., Krewer, C., Selge, C., Müller, F., & Jahn, K. (2016). The subjective postural vertical determined in patients with pusher behavior during standing. Topics in Stroke Rehabilitation, 23(3), 184–190. 10.1080/10749357.2015.1135591

Brötz, D., Johannsen, L., & Karnath, H. O. (2004). Time course of ‘pusher syndrome’ under visual feedback treatment. Physiotherapy Research International, 9(3), 138–143. 10.1002/pri.314

Crawford, J. R., & Garthwaite, P. H. (2005). Testing for suspected impairments and dissociations in single-case studies in neuropsychology: evaluation of alternatives using monte carlo simulations and revised tests for dissociations. Neuropsychology, 19(3), 318–331. 10.1037/0894-4105.19.3.318

Crawford, J. R., & Garthwaite, P. H. (2007). Comparison of a single case to a control or normative sample in neuropsychology: Development of a Bayesian approach. Cognitive Neuropsychology, 24(4), 343–372. 10.1080/02643290701290146

Dai, S., Lemaire, C., Piscicelli, C., & Pérennou, D. (2022). Lateropulsion Prevalence After Stroke: A Systematic Review and Meta-analysis. Neurology, 98(15), e1574–e1584. 10.1212/WNL.0000000000200010

Davies, P. M. (1985). Steps to Follow. A Guide to the Treatment of Adult Hemiplegia (1 ed.). Springer Berlin, Heidelberg.

Johannsen, L., Brötz, D., Nägele, T., & Karnath, H.-O. (2006). " Pusher syndrome" following cortical lesions that spare the thalamus. Journal of Neurology, 253(4), 455–463. 10.1007/s00415-005-0025-7

Johannsen, L., Fruhmann Berger, M., & Karnath, H.-O. (2006). Subjective visual vertical (SVV) determined in a representative sample of 15 patients with pusher syndrome. Journal of Neurology, 253(10), 1367–1369. 10.1007/s00415-006-0216-x

Karnath, H.-O. (2007). Pusher syndrome – a frequent but little-known disturbance of body orientation perception. Journal of Neurology, 254(4), 415–424. 10.1007/s00415-006-0341-6

Karnath, H.-O., & Brötz, D. (2003). Understanding and Treating “Pusher Syndrome”. Physical Therapy, 83(12), 1119–1125. 10.1093/ptj/83.12.1119

Karnath, H.-O., Brötz, D., Baccini, M., Paci, M., & Rinaldi, L. A. (2007). Instructions for the Clinical Scale for Contraversive Pushing (SCP). Neurorehabilitation and neural repair, 21(4), 370–371. 10.1177/1545968307300702

Karnath, H.-O., Brötz, D., & Götz, A. (2001). Klinik, Ursache und Therapie der Pusher-Symptomatik. Der Nervenarzt, 72(2), 86–92. 10.1007/s001150050719

Karnath, H.-O., Ferber, S., & Dichgans, J. (2000a). The neural representation of postural control in humans. Proceedings of the National Academy of Sciences of the United States of America, 97(25), 13931–13936. 10.1073/pnas.240279997

Karnath, H.-O., Ferber, S., & Dichgans, J. (2000b). The origin of contraversive pushing: Evidence for a second graviceptive system in humans. Neurology, 55(9), 1298–1304. 10.1212/WNL.55.9.1298

Karnath, H.-O., Johannsen, L., Brötz, D., & Küker, W. (2005). Posterior thalamic hemorrhage induces “pusher syndrome”. Neurology, 64(6), 1014–1019. 10.1177/0269215509104172

Makowski, D. (2018). The psycho Package: an Efficient and Publishing-Oriented Workflow for Psychological Science. The Journal of Open Source Software, 3(22), 470. 10.21105/joss.00470

Mittelstaedt, H. (1992). Somatic versus vestibular gravity reception in man. Annals of the New York Academy of Sciences, 656, 124–139. 10.1111/j.1749-6632.1992.tb25204.x

Mittelstaedt, H. (1998). Origin and processing of postural information. Neuroscience & Biobehavioral Reviews, 22(4), 473–478. 10.1016/S0149-7634(97)00032-8

Nestmann, S., Röhrig, L., Müller, B., Ilg, W., & Karnath, H.-O. (2022). Tilted 3D visual scenes influence lateropulsion: A single case study of pusher syndrome. Journal of Clinical and Experimental Neuropsychology, 44(7), 478–486. 10.1080/13803395.2022.2121382

Pedersen, P. M., Wandel, A., Jørgensen, H. S., Nakayama, H., Raaschou, H. O., & Olsen, T. S. (1996). Ipsilateral pushing in stroke: Incidence, relation to neuropsychological symptoms, and impact on rehabilitation. The Copenhagen stroke study. Archives of Physical Medicine and Rehabilitation, 77(1), 25–28. 10.1016/S0003-9993(96)90215-4

Posit Software. (2024). RStudio: Integrated Development Environment for R. In (Version 4.4.0) Posit Software, PBC. http://www.posit.co/.

Rittmo, J. Ö., & McIntosh, R. D. (2021). singcar: Comparing single cases to small samples in R. Journal of Open Source Software, 6(68), 3887. 10.21105/joss.03887

Rosenzopf, H., Klingbeil, J., Wawrzyniak, M., Röhrig, L., Sperber, C., Saur, D., & Karnath, H.-O. (2023). Thalamocortical disconnection involved in pusher syndrome. Brain, 00, 1–14. 10.1093/brain/awad096

van der Waal, C., Embrechts, E., Loureiro-Chaves, R., Gebruers, N., Truijen, S., & Saeys, W. (2022). Lateropulsion with active pushing in stroke patients: its link with lesion location and the perception of verticality. A systematic review. Topics in Stroke Rehabilitation, 1–17. 10.1080/10749357.2022.2026563

Wöhrstein, S., Bressler, M., Röhrig, L., Prahm, C., & Karnath, H.-O. (2025). A head-mounted Tilted Reality Device for the treatment of pusher syndrome: a usability study in healthy young and older adults. Virtual Reality, 29(1), 2. 10.1007/s10055-024-01066-0

Yang, Y.-R., Chen, Y.-H., Chang, H.-C., Chan, R.-C., Wei, S.-H., & Wang, R.-Y. (2015). Effects of interactive visual feedback training on post-stroke pusher syndrome: a pilot randomized controlled study. Clinical Rehabilitation, 29(10), 987–993. 10.1177/0269215514564898

